# Epidemiological Surveillance of Amyotrophic Lateral Sclerosis: A Review

**DOI:** 10.1101/2023.11.10.23297968

**Authors:** Christina Wolfson, Danielle E. Gauvin, Foluso Ishola, Maryam Oskoui, Boris Atabe

**Affiliations:** Neuroepidemiology Research Unit, Research Institute of the McGill University Health Centre, Montreal, Quebec, Canada; Department of Medicine, Faculty of Medicine and Health Sciences, McGill University, Montreal, Quebec, Canada; Department of Epidemiology, Biostatistics and Occupational Health, School of Population and Global Health, McGill University, Montreal, Quebec, Canada; Department of Pediatrics, Neurology & Neurosurgery, Faculty of Medicine and Health Sciences, McGill University, Montreal, Quebec, Canada

**Keywords:** Amyotrophic Lateral Sclerosis, Surveillance, Registries, Epidemiology

## Abstract

**Background:** Registries and clinical databases are important tools to systematically record and collect information about individuals with rare diseases and to monitor disease patterns in populations. Through a review of the published literature on strategies used for surveillance of Amyotrophic Lateral Sclerosis (ALS), our objective was to better delineate the varied approaches used to monitor ALS at a population level. Further, we sought to determine the potential of registries to enhance knowledge on the epidemiology of ALS using a case study comparing epidemiological outputs from registries in the United States, United Kingdom, and Italy.

**Summary:** We searched Medline, Embase, Global Health, PsycInfo, Cochrane Library, and CINAHL identifying articles published between January 1^st^, 2010, and May 12^th^, 2021. Studies describing population registries, cohorts of individuals with ALS, or large-scale studies aimed at systematically identifying people with ALS, were eligible for inclusion. 1,447 publications were found, of which 141 were selected for full text review, and 41 of those were selected for data extraction. We identified ALS registries and pertinent databases in 4 continents (North America, Europe, Asia, and Oceania). Stated objectives of the registries/databases shaped their framework, methodology, and follow-up. The US National Registry demonstrates substantial research outputs and methodological strengths, producing many descriptive epidemiological outputs (n=5 studies) and several methodological papers (n=12 studies). The UK and Italy overall each produced a similar number of studies (albeit with fewer methodological papers), across several different registries and regions.

**Key Messages:** Due to challenges inherent to the surveillance of rare diseases, registries are a vital tool in determining and assessing the global impact of ALS. Nevertheless, the development and implementation of registries is not feasible everywhere in the world. There are advantages and drawbacks to structuring registries at a national or regional level, often dictated by funding availability, resources and health care infrastructure, and research objectives. To fully assess the epidemiological burden of ALS globally, collaborative initiatives are needed to fill gaps in knowledge, and there is a critical need to harmonize and optimize the development, collection, and sharing of data across registries.

## Introduction

Amyotrophic Lateral Sclerosis (ALS) is a rare and rapidly fatal neurodegenerative motor neuron disease (MND), affecting upper and lower motor neurons. These characteristics, along with challenges in clinical diagnosis in regions with limited neurological resources and health care accessibility, make it difficult, if not impossible in some settings, to conduct population-based epidemiological research in ALS. Challenges in surveillance of rare diseases, that is, capturing and collecting data on individuals with a rare disease at a population level, also make it difficult to assess the true impact of ALS on a larger, global scale. Several research groups around the world have established patient registries or clinical databases to systematically identify and collect information about individuals with ALS to facilitate research and to monitor disease patterns in the population. There are several definitions for a registry and for the purposes of this paper, we use the definition used by Arts et al. (2002) “*a systematic collection of a clearly defined set of health and demographic data for patients with specific health characteristics, held in a central database for a predefined purpose.*” [1] This definition captures important principles of a registry, that is, information is collected systematically, clearly defined, held centrally, and for a pre-defined purpose. Some studies outlined in this paper employ the terminology of a register, while others use registry. For consistency, we refer to all of these as registries.

A systematic review by Barbalho et al. (2021) presents guidelines for structuring population-based registries of motor neuron diseases, detailing aspects of registry development for five registries (two in Europe, three in North America, and one in Oceania). [2] Their work underscores important, diverse, and robust strategies for information capture, data access, and emphasizes the need for patient-focused registries, that is, registries that are motivated by and developed with, the aim of furthering patient care and treatment, in addition to furthering research objectives, including clinical trials. Here, we aim to add to this ongoing conversation by detailing research that attempts to systematically collect information on individuals with ALS, not limiting design/methodology to a registry, specifically.

To better understand the strategies used to monitor ALS at a population level, the primary objective of this review was to examine the published literature to identify and describe existing methods of ALS surveillance anywhere in the world. Secondarily, we aimed to assess if ALS registries enhance research and knowledge on the epidemiology of ALS by comparing the volume of published research produced from ALS registries in three different regions (United States, United Kingdom, and Italy).

## Methods

### Search strategy

We searched Medline, Embase, Global Health, PsycInfo, Cochrane Library, and CINAHL to identify relevant articles published between January 1st, 2010, and May 12th, 2021. The search strategy was developed in collaboration with a librarian (AB) through the McGill University Health Centre Library (see Supplemental Materials). No language restrictions were imposed in the initial search. This study is registered with PROSPERO under ID number CRD42021250572. The authors also ran, in parallel, a separate search on the global epidemiology of ALS (PROSPERO ID number CRD42021250559) which is the subject of another manuscript [3]. Studies deemed relevant to the surveillance review were flagged during the abstract and title screening of the global epidemiology review and included (if not already) in this review. This strategy added only one publication.

### Inclusion and exclusion criteria

Inclusion criteria required that publications be studies of epidemiological monitoring, surveillance, and/or disease registries of ALS. This included articles that describe population registries, large cohorts of individuals with ALS, or studies that attempted to systematically identify large groups of individuals with ALS. Systematic reviews and meta-analyses were excluded but their references were searched. Any duplicate publications, conference proceedings, case reports, and/or clinical trials were excluded. Articles with an abstract only, where no full text was available, and articles published in a language other than French or English for which translation was not possible were also excluded. We did not include registries that were derived exclusively from patient self-reports.

### Quality appraisal

Since the goal of this review was to identify and describe ALS surveillance and monitoring strategies, we did not conduct quality assessment of the selected publications. Overall strengths and limitations for each individual registry are addressed in this review.

### Data extraction

Covidence software was used to manage this review. Screening of abstracts for eligibility was completed by three reviewers (DG, FI, CW) independently. Articles at each stage of the review process required two reviewers to vote, while the third adjudicated disagreements. After initial title and abstract screening, full-text versions of the selected articles were obtained and further reviewed to determine eligibility and inclusion in the review. Again, this was completed by two reviewers, with disagreements adjudicated by the third reviewer. Finally, two reviewers independently assessed all eligible studies and compiled relevant information in a data extraction form. As our main outcomes were methodological approaches for the monitoring and surveillance of ALS, the following information was sought: study parameters (title, author, publication year, region and country, study design, registry name, registry launch year, registry objective(s), study period and duration, and disease(s) included in the registry/study), study population characteristics (catchment area, size of study population, age, ethnicity, sex, representativeness of target population), case definition of ALS (case ascertainment method(s), diagnostic method(s), data source(s)), methodology (study limitations and potential biases, patient consent, funding, maintenance of registry, and users of the registry), and information collected as part of the registry (under the following categories: sociodemographic, general health, clinical data, respiratory status, medication, cognition, functional status, quality of life, health care utilization, and other). The data extraction form was piloted on selected studies by three team members (DG, FI, CW).

### Data synthesis

Our synthesis of the results describes the geographical distribution of ALS registries, which population(s) are included and how ALS is defined, how cases are captured (case ascertainment was categorized into active, passive, linkage), what kind of information is collected, data sources, and considerations/barriers in the implementation of registries. Data visualization was performed using R version 4.1.0 [4] and mapchart.net.

### Case study

To assess whether ALS registries, as a surveillance platform, enhance research and knowledge on the epidemiology of ALS, we compare and contrast three regions with differing ALS registry landscapes: United States (a single national registry), United Kingdom (a mixture of six national and regional registries), and Italy (five regional registries). Relevant studies from the aforementioned epidemiology systematic review were a starting point for registry outputs (incidence, prevalence, mortality estimates), and references of these articles were searched for any publications describing registry methods in detail. An additional search (PubMed and Medline) of the literature was carried out in July 2022 on these registries to collate direct outputs using registry data, that is, how the registry was/is being used for epidemiological purposes. This allowed for the inclusion of methodological papers outside the initial review time frame, and recent studies that utilized registry data for epidemiological research (i.e., articles published before 2010, and those published more recently in 2022). Inclusion criteria for this additional search were: population-based observational study describing the selected registries, or studies that had utilized registry data to estimate ALS incidence, prevalence, or mortality. Further, the article must include a statement of the primary objective of the registry and how cases for the registry were ascertained and recorded.

### Data availability

Requests for access to the data reported in this article will be considered by the corresponding author.

## Results

A flowchart of the study selection process is presented in Figure 1. From all database searches, 1,447 articles were included for title and abstract screening, after removal of duplicates. Following title and abstract screening, 141 articles were selected for full text review. Following full text review, a total of 41 articles were selected for data extraction. We identified ALS registries and pertinent databases from four continents (North America, Europe, Asia, and Oceania) and within these continents, 15 countries (USA, Canada, Germany, Italy, the Netherlands, Sweden, Norway, Spain, France, Scotland, Iran, China, South Korea, Australia, and New Zealand). The geographical distribution of registries is shown in Figure 2. In our search we did not find any registries or comprehensive databases that met our inclusion criteria from Africa or South America. Table S1 includes the stated objectives of each registry or database. Generally, the main objective(s) related to the creation of a platform for research on the incidence and/or prevalence of, and survival/mortality of ALS. Several of the registries were initially developed to enhance patient access and recruitment for clinical trials of treatment and/or to coordinate clinical care. Over time, these then became sources of information used in epidemiological research.

**Figure 1.**
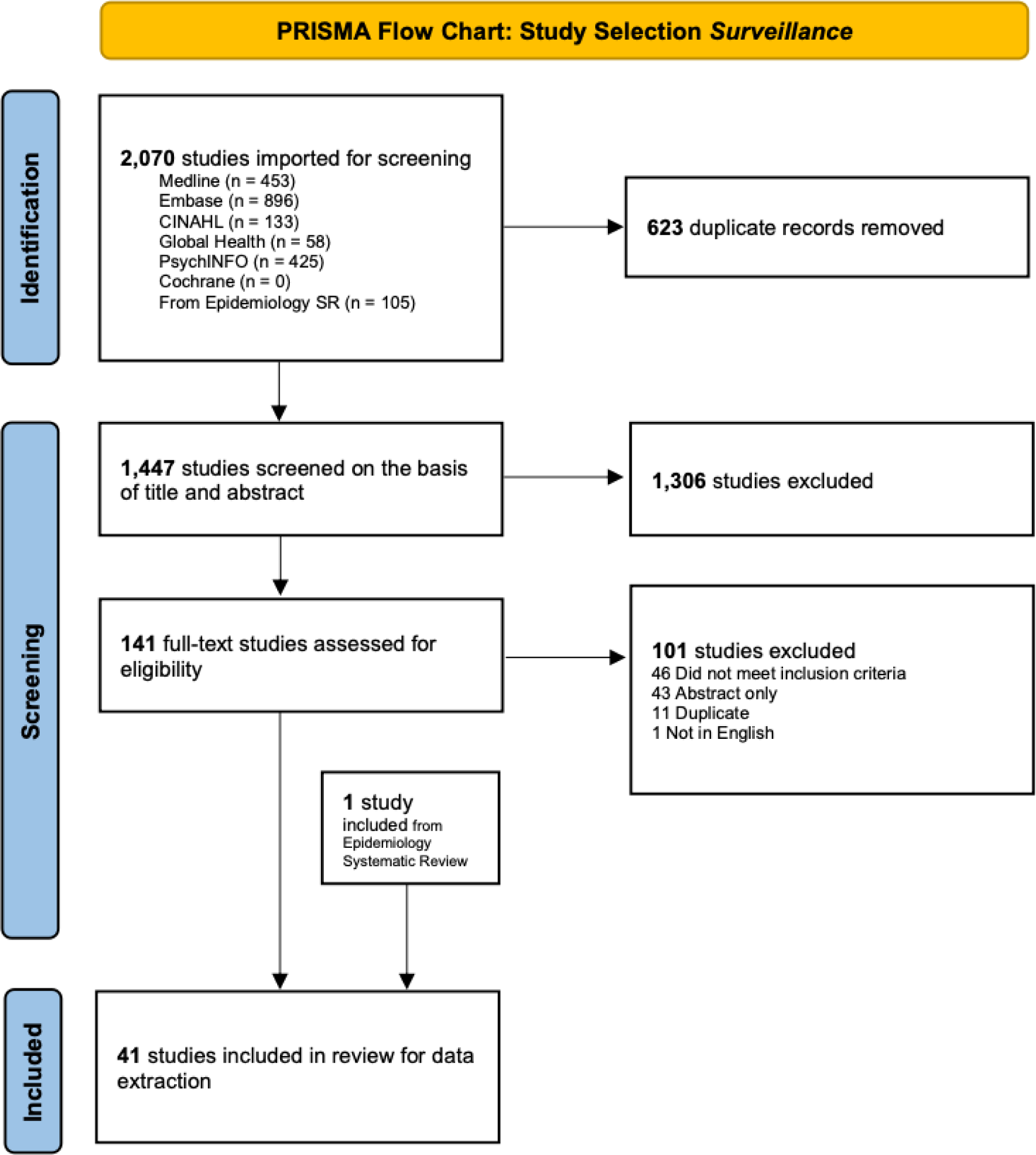
PRISMA study selection flowchart.

**Figure 2.**
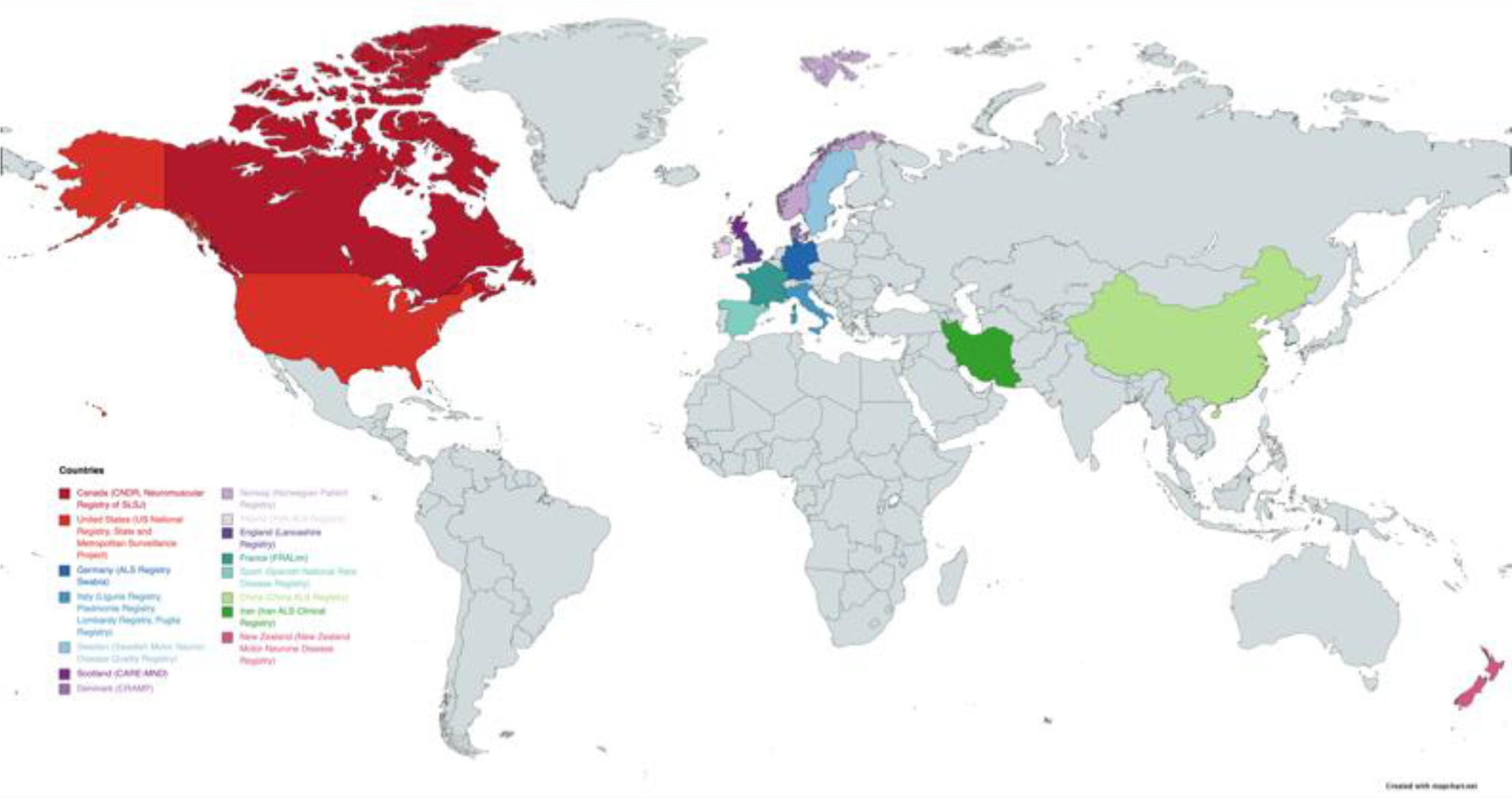
Geographical distribution of ALS/MND registries.

### Methodology of registries/databases

Table 1 provides an overview of the methodology of each registry and database. Launch year ranged from 1985 (Neuromuscular Registry of Saguenay-Lac-Saint-Jean, SLSJ [5]) to 2020 (China ALS Registry, CHALSR [6]). Overall, common data sources included neurology clinics and administrative data (hospital and death records) with many registries incorporating data from multiple sources. The New Zealand Motor Neurone Disease Registry [7] incorporates data from community ALS organizations (e.g., membership lists). A few of the surveillance projects in the United States (Massachusetts Tracking Program [8] and the Integrated Neurodegenerative Disease Database [9]) utilized data from existing patient registries. The National ALS Registry [10-19] in the USA also integrated data from the Veterans’ Health Administration (VHA), Veterans’ Benefit Administration (VBA), Medicare, and Medicaid.

**Table 1.**
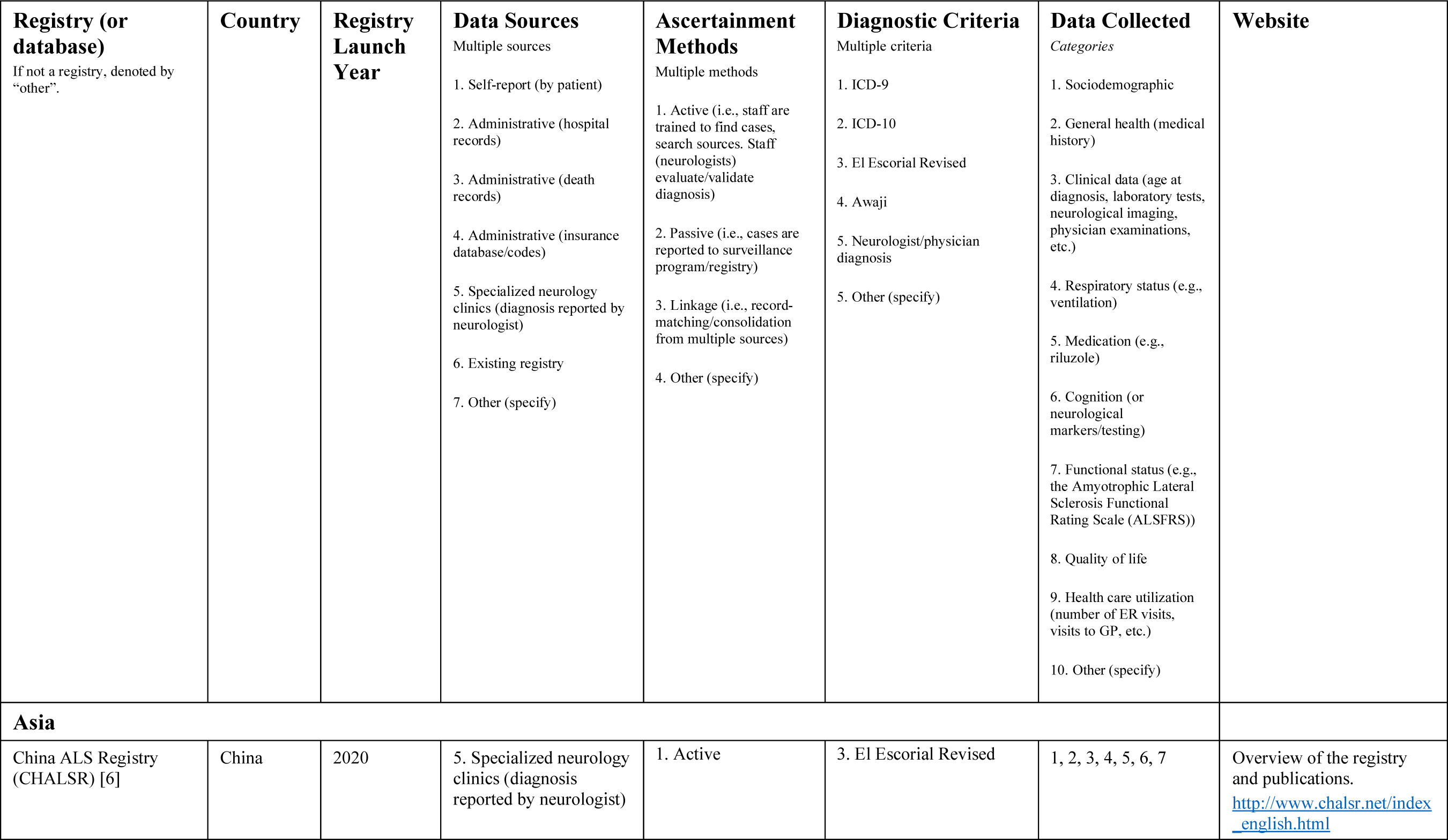
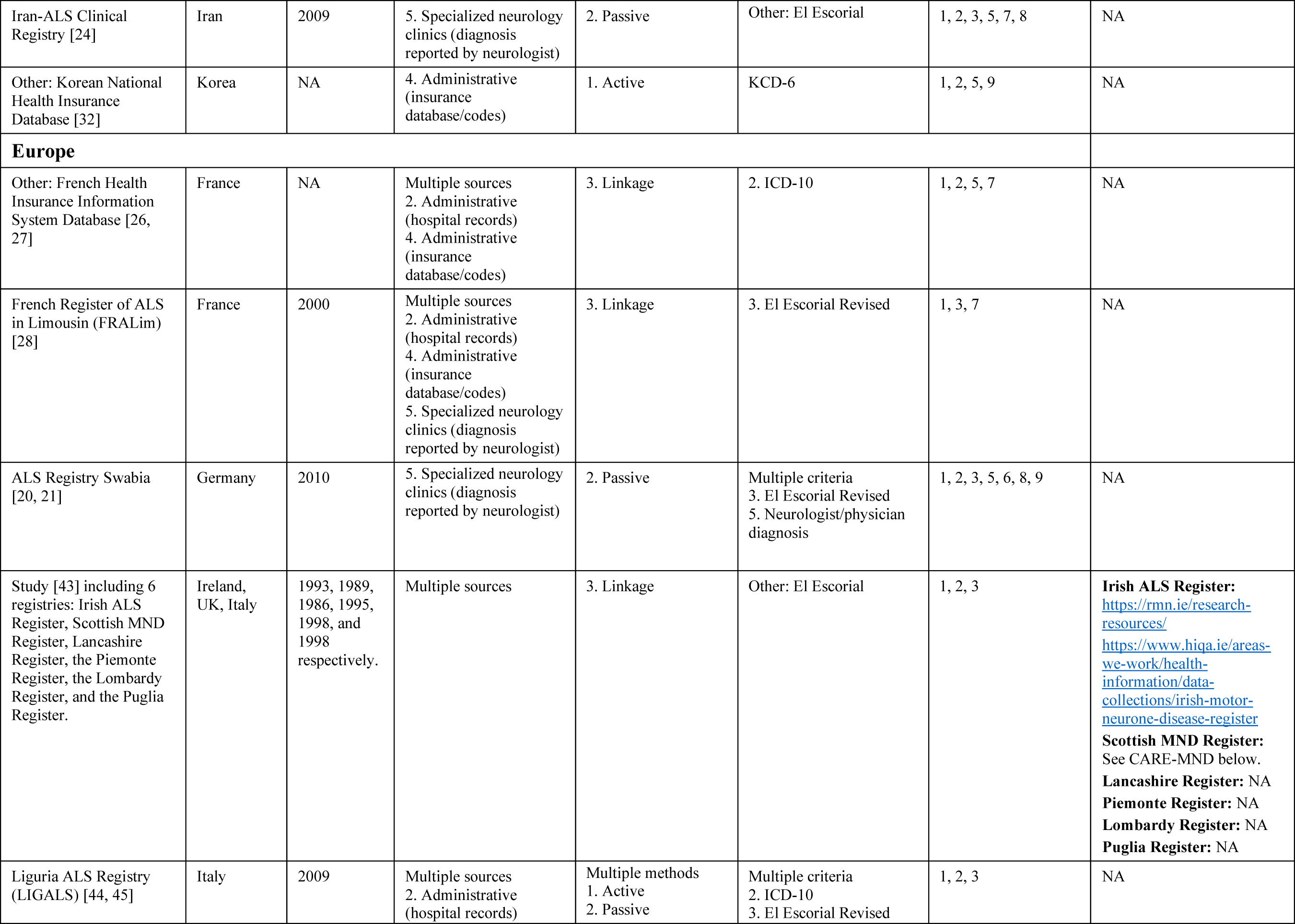
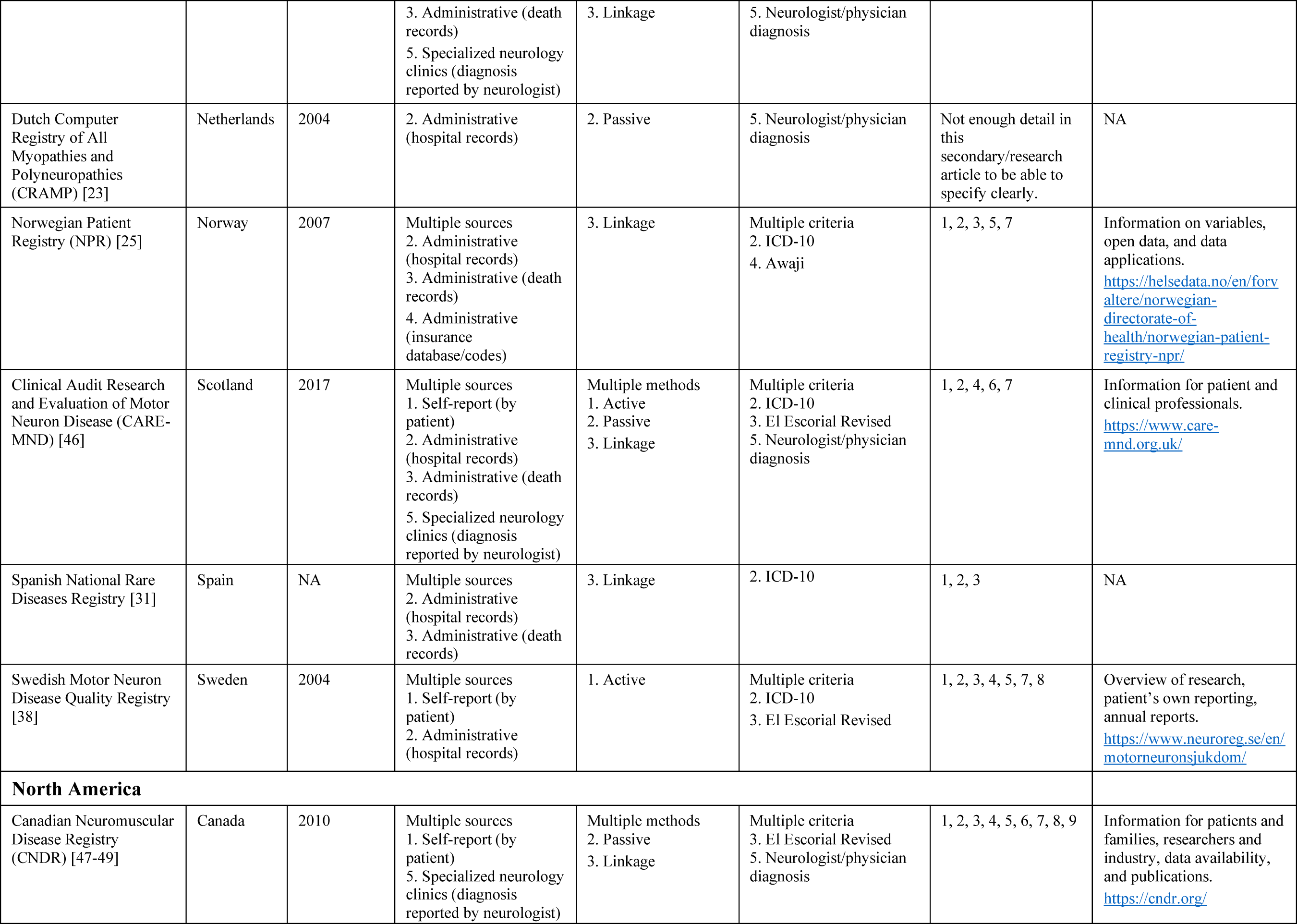
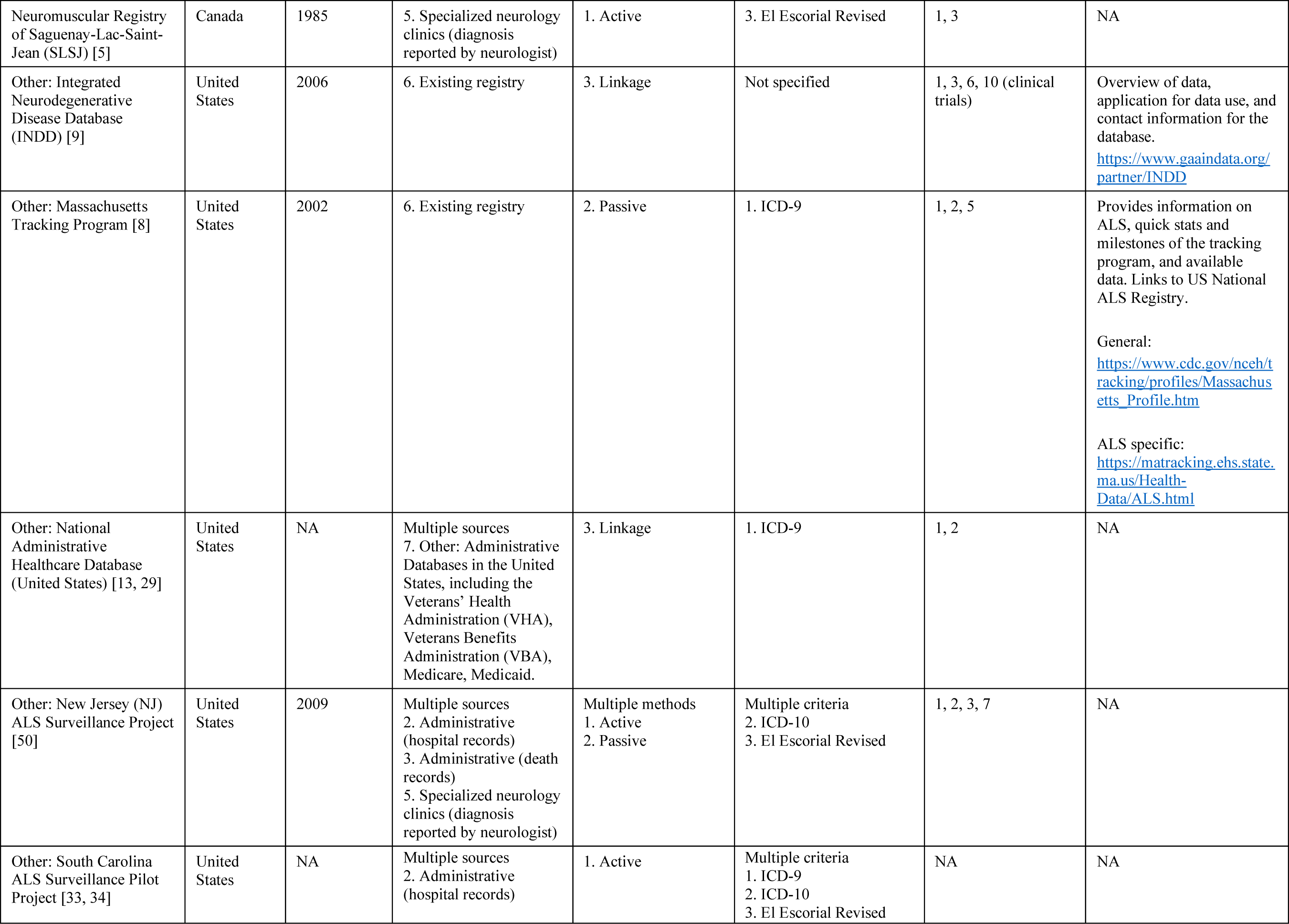
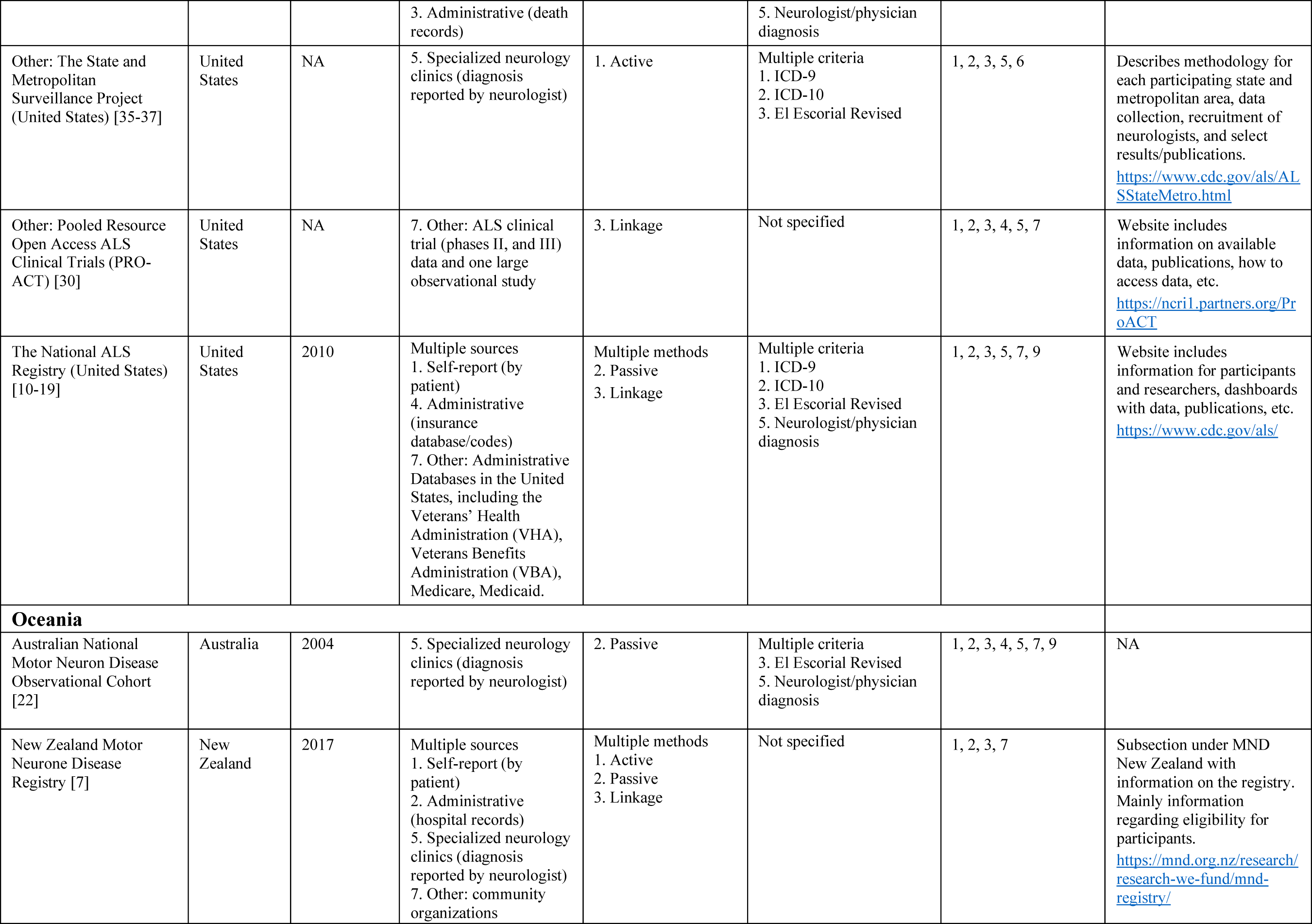
General overview of the methodology of registries. We report country, registry launch year (if applicable), data sources (where cases are taken from), ascertainment methods (how cases are collected), diagnostic criteria, and data collected as part of the registry (information available).

Many registries rely only on passive (i.e., voluntary reporting of cases directly to the surveillance entity, by physicians and/or through self-report) methods to capture individuals with ALS. This is the case for ALS Registry Swabia in Germany [20, 21], the Australian National Motor Neuron Disease Observational Cohort [22], the Dutch Computer Registry of All Myopathies and Polyneuropathies (CRAMP) [23], the Iran ALS Clinical Registry [24], and the Massachusetts (USA) Tracking Program [8]. Linkage (record-matching, consolidation of cases from many sources) was the main method of case ascertainment for the Norwegian Patient Registry (NPR) [25], the French Health Insurance Information System Database [26, 27], the French Register of ALS in Limousin (FRALim) [28], the Integrated Neurodegenerative Disease Database (INDD) [9] in the United States, the National Administrative Healthcare Database (USA) [13, 29], the Pooled Resource Open Access ALS Clinical Trials (PRO-ACT) [30] in the USA, and the Spanish National Rare Diseases Registry [31]. Active recruitment of cases, where multiple sources are searched and staff are specifically trained to find cases, was the main method of ascertainment in the China ALS Registry (CHALSR) [6], SLSJ [5], Korean National Health Insurance Database [32], South Carolina ALS Surveillance Project [33, 34], State and Metropolitan Surveillance Project (United States) [35-37], and the Swedish Motor Neuron Disease Quality Registry [38]. Many registries use multiple methods of case ascertainment. Registry creators rely on resources available to them for the systematic collection of cases in line with standards in diagnostic criteria. Most registries used the El Escorial Criteria (Revised or Original) as diagnostic criteria, with most supplementing this with a diagnosis reviewed and validated by a neurologist and/or ICD-9 or ICD-10 codes for MND/ALS.

At a minimum, all registries and databases, collected information on sociodemographic characteristics (age, sex), health (e.g., medical history), and/or clinical data (e.g., age at diagnosis, neurological imaging, etc.) for each patient. Other information commonly included was medication (e.g., riluzole prescription), functional status (e.g., ALS Functional Rating Scale [39]), and health care utilization (e.g., number of emergency room visits, hospital discharge data, etc.). One must also take into consideration the additional resources needed to collect longitudinal data on individual patients entered into the registry beyond initial intake information. Ongoing inclusion of new patients and having the proper mechanisms/resources in place to maintain patient records through follow-up of individuals from entry to death or loss to follow-up is vital for the success of an ALS registry.

### Do ALS registries enhance research and knowledge on the epidemiology of ALS? A case study

In the United States, the National ALS Registry (launch year 2010) was designed and implemented to identify ALS cases throughout the entire country (open to all US citizens or residents with ALS). Registries in the UK included the Northern Ireland ALS Register (launched in 1993), the Irish ALS Register (launched in 1993), the Scottish MDN Register (launched in 1989, relaunched as CARE-MND in 2015), the Lancashire Register (launched in 1986), the South East ALS Register (SEALS; launched in 2002), and the MND Register for England, Wales, and Northern Ireland (launched in 2002). In Italy, the registries were: the Piemonte and Valle d’Aosta Register for ALS (PARALS; launched in 1995), the Sclerosi Laterale Amiotrofica Puglia Register (SLAP; launched in 1997), SLA Lombardy (SLALOM; launched in 1998), the Emilia Romagna Registry (ERRALS; launched in 2009), and the Liguria Amyotrophic Lateral Sclerosis Registry (LIGALS; launched in 2009). Figure 3 displays the timeline of published studies from each region. The US National Registry had the most outputs (and the most recent, between 2010 and 2020); notably, prevalence estimates are updated and published on a yearly basis (n = 5 descriptive epidemiology studies). Detailed methodology is available across multiple studies (n = 12) for the US National ALS Registry, this included pilot projects and validation studies, namely the State and Metropolitan Surveillance Project carried out over several cities and states across the US. These were conducted to evaluate the completeness and reliability of incidence/prevalence estimates of the US National Registry, and further describe the distributions of age, sex, and race (among other characteristics) of individuals living with ALS in the USA. In the UK, each regional registry was produced a moderate number of outputs from the mid-1990s to 2021 (at least two descriptive epidemiology studies per registry, except for the MND Register for England, Wales, and Northern Ireland for which we only found one descriptive study), there were few dedicated studies which described registry methodology (n = 2, both for CARE-MND). Comparatively, Italy had lot of regional/local registries, but with limited outputs for each (we found one descriptive epidemiology study each for SLALOM and SLAP, two for LIGALS, while ERRALS and PARALS both had n = 3 studies). Publication year ranging from the early 2000s to 2022. Our search yielded only one methodological papers for the Italian registries (SLALOM [40]). It is worth noting, given the smaller catchment areas in regional registries, with such a rare disease the smaller number of cases likely reduces the opportunity to publish robust population estimates. It is of note that we do not have any information on submitted publications that were not published or on manuscripts submitted and under review at the time of our review.

**Figure 3.**
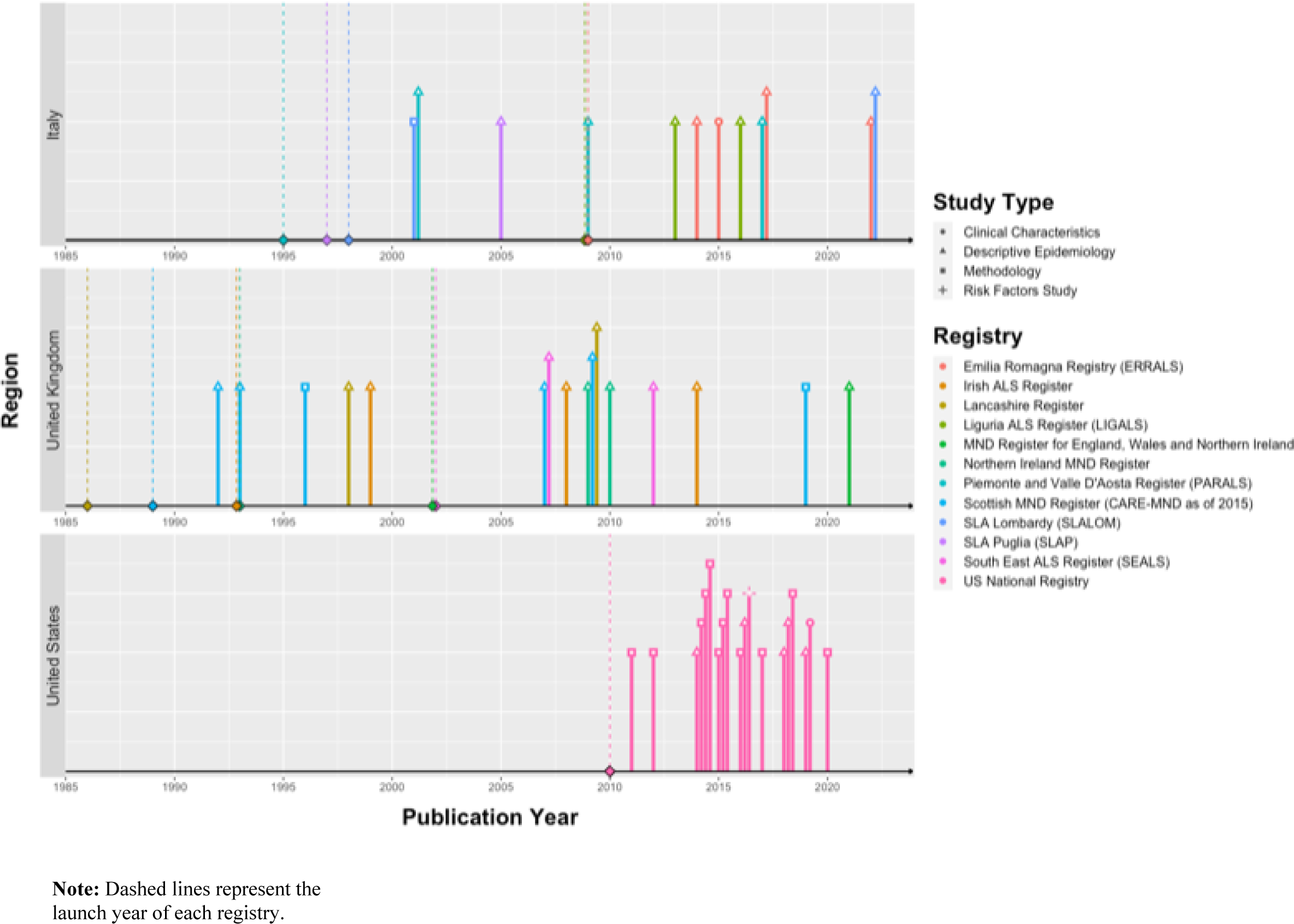
Case Study: Timeline of published studies from registries in Italy, the United Kingdom, and the United States.

## Conclusion

Registries can enhance comparisons across regions/countries and harmonization of data. We identified ALS registries and pertinent databases in North America, Europe, Asia and Oceania. The objectives of these registries/surveillance strategies guided their structure, methodology, and follow-up. Some registries were based on linkage with administrative health or insurance databases. It is resource intensive for developers of registries to conduct a review of each possible case of ALS for inclusion in the registry. For most registries, cases were obtained from multiple sources. Identifying and harmonizing these sources also requires resources, and concerns of under-ascertainment (i.e., that not all cases may be fully captured by available data) require additional steps which have included the application of capture-recapture methodology to estimate the number of missing cases when multiple sources are used. This methodology is only useful in adjusting estimates of prevalence and incidence; but does not provide individual data on these unascertained cases. Not all registries required participant consent and this review did not contrast consent-based registries vs. those that do not require patient consent. It is possible that individuals with ALS who choose to not participate in a registry are different compared to individuals included in a registry, in terms of clinical characteristics. This may lead to selection bias, affecting the accuracy and generalizability of survival and mortality estimates and bias in other estimated epidemiological parameters (e.g., relative risk). Certainly, with a disease as rare as ALS, even a small number of missing cases in a relatively small population could have a marked influence on incidence and prevalence.

Given the methodological strengths (e.g., patient recruitment, case ascertainment, registry maintenance) and substantial number of outputs (i.e., studies that utilized registry data), the authors view the US National Registry as an important model for the development and implementation of an ALS registry. Collecting and storing data in one large central entity allows consistent methodology and follow-up of a large number of cases. The US National Registry also routinely updates and publishes estimates of national incidence and prevalence of ALS, in line with their registry objectives. It is worth noting that this is largely possible due to adequate sustained funding, infrastructure, and expertise. There are also considerations such as the collaborative involvement of hospital networks and local clinics to ensure sufficient coverage for patient recruitment, neurologists that can validate clinical criteria for diagnosis, and resources dedicated to databases that house, maintain, and update data, securely. [2] These are luxuries that are not afforded to researchers everywhere in the world. Nevertheless, there is value in the development and implementation of registries in smaller geographic areas. Smaller local registries allow for regional comparisons of incidence and prevalence within a country which is important given that disease burden may differ within a country. This is especially pertinent given barriers in accessing care, particularly amongst marginalized populations. On an individual level, earlier diagnosis enables earlier access to interdisciplinary care and management of symptoms. [41, 42] One might suggest that regional registries, while limited in the number of epidemiological outputs (i.e., number of studies), may directly inform patient-centered approaches to care and resource allocation even in the absence of peer review published research. Not only are registries a large endeavour to develop and establish, but they also require a lot of time and resources to maintain. Scaling smaller, in terms of registry design, may alleviate some of these burdens while capturing important region-specific information that can be used to directly improve community access to care and treatment. However, this is directly dependent on expertise (i.e., local researchers/neurologists) and funding, which can be challenging to assemble in remote and/or under resourced areas.

This review sought to identify published information on registries and systematic collections of individuals with ALS worldwide. There were many differences not only in the objectives in the various surveillance infrastructures identified, but also in the methods used. With the hope that registries will over time enhance the ability to conduct epidemiological research, in particular to monitor incidence and prevalence, there is a need to harmonize and optimize the development of, and collection of data in, registries.

## Supporting information

Search Strategy

## Acknowledgements

The authors gratefully acknowledge funding from the Public Health Agency of Canada which enabled this work. The views expressed herein do not necessarily represent the views of the Public Health Agency of Canada. In terms of developing and fine tuning the literature searches, we thank Amy Bergeron, McGill University Health Centre Library for their guidance and expertise. We thank Karen Zabowski for her assistance in the financial and human resources aspects of the project. Boris Atabe was awarded a summer studentship from the BRaIN Repair and Integrative Neuroscience Program at the Research Institute of the McGill University Health Center and the International Brain Research Organization. In closing, we acknowledge the millions of people living with Amyotrophic Lateral Sclerosis around the world and their loved ones.

## Author Contributions

CW was responsible for conceptualization, funding, project administration, methodology, investigation, analysis and interpretation of data, validation, and drafting of the manuscript. DG was responsible for conceptualization, data curation, project administration, methodology, investigation, analysis and interpretation of data, validation, and drafting of the manuscript. MO was responsible for methodology, investigation, validation, and editing of the manuscript. FI was responsible for methodology, investigation, validation, and editing of the manuscript. BA was responsible for the methodology and investigation of the case study portion of this work. All authors had full access to all the data in the study, verified the data, and had final responsibility for the decision to submit for publication.

