## Supplementary material for "Epidemiological Surveillance of Amyotrophic Lateral Sclerosis: A Review": Search Strategy

**Medline**

1. Amyotrophic Lateral Sclerosis/ or Motor Neuron Disease/
2. (Amyotrophic Lateral Sclerosis or motor neuron* disease* or Lou Gehrig* or Charcot* disease*).ti,ab,kf.
3. 1 or 2
4. Population Surveillance/ or Public Health Surveillance/ or Sentinel Surveillance/ or Epidemiological Methods/ or Epidemiological Monitoring/
5. (((disease*) adj2 (detect* or report*)) or ((surveill*) adj4 (method* or strateg* or approach* or system* or population* or health* or sentinel* or disease*))).ti,ab,kf.
6. ((disease* or epidemiolog* or public health) adj3 (monit*)).ti,ab,kf.
7. (((surveill* or monitor* or report* or detect*) adj6 (public health or registr*)) or ((disease* or prov* or health) adj2 (registr* or database)) or registry or registries).ti,ab,kf.
8. 4 or 5 or 6 or 7
9. 3 and 8
10. limit 9 to yr="2010-Current"

**Embase**

1. Amyotrophic Lateral Sclerosis/ or Motor Neuron Disease/
2. (Amyotrophic Lateral Sclerosis or motor neuron* disease* or Lou Gehrig* or Charcot* disease*).ti,ab,kw.
3. 1 or 2
4. Health Survey/ or Sentinel Surveillance/ or Epidemiological Monitoring/
5. (((disease*) adj2 (detect* or report*)) or ((surveill*) adj4 (method* or strateg* or approach* or system* or population* or health* or sentinel* or disease*))).ti,ab,kw.
6. ((disease* or epidemiolog* or public health) adj3 (monit*)).ti,ab,kw.
7. (((surveill* or monitor* or report* or detect*) adj6 (public health or registr*)) or ((disease* or prov* or health) adj2 (registr* or database)) or registry or registries).ti,ab,kw.
8. 4 or 5 or 6 or 7
9. 3 and 8
10. limit 9 to yr="2010-Current"

**CINAHL**

1. (MH "Amyotrophic Lateral Sclerosis") or (MH "Motor Neuron Disease")
2. TI (Amyotrophic Lateral Sclerosis or motor neuron* disease* or Lou Gehrig* or Charcot* disease*) or AB (Amyotrophic Lateral Sclerosis or motor neuron* disease* or Lou Gehrig* or Charcot* disease*)
3. 1 or 2

4. (MH "Disease Surveillance") or (MH "Population Surveillance") or (MH "Registries, Disease")
5. TI (((disease*) N2 (detect* or report*)) or ((surveill*) N4 (method* or strateg* or approach* or system* or population* or health* or sentinel* or disease*))) or AB (((disease*) N2 (detect* or report*)) or ((surveill*) N4 (method* or strateg* or approach* or system* or population* or health* or sentinel* or disease*)))
6. TI ((disease* or epidemiolog* or public health) N3 (monit*)) or AB ((disease* or epidemiolog* or public health) N3 (monit*))
7. TI (((surveill* or monitor* or report* or detect*) N6 (public health or registr*)) or ((disease* or prov* or health) N2 (registr* or database)) or registry or registries) or AB (((surveill* or monitor* or report* or detect*) N6 (public health or registr*)) or ((disease* or prov* or health) N2 (registr* or database)) or registry or registries)
8. 4 or 5 or 6 or 7
9. 3 and 8

**<u>Global Health</u>**

1. (Amyotrophic Lateral Sclerosis and ALS).ti,ab,hw.
2. (Amyotrophic Lateral Sclerosis or motor neuron* disease* or Lou Gehrig* or Charcot* disease*).ti,ab,hw.
3. 1 or 2
4. Surveillance/ or Sentinel Surveillance/ or Epidemiological Surveys/ or Monitoring/ or Data Collection/
5. (((disease*) adj2 (detect* or report*)) or ((surveill*) adj4 (method* or strateg* or approach* or system* or population* or health* or sentinel* or disease*))).ti,ab,hw.
6. ((disease* or epidemiolog* or public health) adj3 (monit*)).ti,ab,hw.
7. (((surveill* or monitor* or report* or detect*) adj6 (public health or registr*)) or ((disease* or prov* or health) adj2 (registr* or database)) or registry or registries).ti,ab,hw.
8. 4 or 5 or 6 or 7
9. 3 and 8
10. limit 9 to yr="2010-Current"

**<u>PsychINFO</u>**

1. Amyotrophic Lateral Sclerosis/
2. (Amyotrophic Lateral Sclerosis or motor neuron* disease* or Lou Gehrig* or Charcot* disease*).ti,ab,id.
3. 1 or 2
4. Monitoring/
5. (((disease*) adj2 (detect* or report*)) or ((surveill*) adj4 (method* or strateg* or approach* or system* or population* or health* or sentinel* or disease*))).ti,ab,id.

6. ((disease* or epidemiolog* or public health) adj3 (monit*)).ti,ab,id.
7. (((surveill* or monitor* or report* or detect*) adj6 (public health or registr*)) or ((disease* or prov* or health) adj2 (registr* or database)) or registry or registries).ti,ab,id.
8. 4 or 5 or 6 or 7
9. 3 and 8
10. limit 9 to yr="2010-Current"

**Cochrane**

1. Amyotrophic Lateral Sclerosis/ or Motor Neuron Disease/
2. (Amyotrophic Lateral Sclerosis or motor neuron* disease* or Lou Gehrig* or Charcot* disease*).ti,ab,kf.
3. 1 or 2
4. Population Surveillance/ or Public Health Surveillance/ or Sentinel Surveillance/ or Epidemiological Methods/ or Epidemiological Monitoring/
5. (((disease*) NEAR/2 (detect* or report*)) or ((surveill*) NEAR/4 (method* or strateg* or approach* or system* or population* or health* or sentinel* or disease*))).ti,ab,kf.
6. ((disease* or epidemiolog* or public health) NEAR/3 (monit*)).ti,ab,kf.
7. (((surveill* or monitor* or report* or detect*) NEAR/6 (public health or registr*)) OR ((disease* or prov* or health) NEAR/2 (registr* or database)) or registry or registries).ti,ab,kf.
8. 4 or 5 or 6 or 7
9. 3 and 8
