## Supplemental Table S1 for "Epidemiological Surveillance of Amyotrophic Lateral Sclerosis: A Review"

**Table S1.** List of registries stated objectives, and diseases/disorders included.

| Registry | Stated Objective(s)<br>Extracted directly from articles | Disease(s) Included |
|---|---|---|
| **Asia** | | |
| China ALS Registry (CHALSR) [e1] | The aim of CHALSR is "to explore the natural history and clinical features of ALS in mainland China through a multicentre, prospective cohort. The specific objectives include the following: 1) Investigate the geographical and temporal distribution of ALS patients in mainland China from 2020 to 2023. 2) Describe the clinical characteristics and disease progression of ALS. 3) Identify the factors that impact the prognosis of ALS." [e1(p.2)] | ALS |
| Iran-ALS Clinical Registry[e2] | The main aim was "to evaluate one-year disease progression using a physical and functional disability scoring system in Iranian patients through a prospective multicenter study. The second goal was to compare the effects of riluzole on disease progression." [e2(p.506)] | ALS |
| Other: Korean National Health Insurance Database[e3] | This study "aimed to determine the incidence, prevalence and survival time of Korean patients with amyotrophic lateral sclerosis (ALS) using National Health Insurance Service (NHIS) data." [e3(p.395)] | ALS |
| **Europe** | | |
| Other: French Health Insurance Information System Database[e4,5] | Use the French Health Insurance Information System Database linked to the French Hospital Discharge Database to identify incident MND cases in France, and to compare. | MND |
| French Register of ALS in Limousin (FRALim) [e6] | "The main objective of establishing the French register of amyotrophic lateral sclerosis (ALS) in the Limousin region (FRALim), was to assess the incidence of ALS, in this ageing region of Europe, over a 12-year period (2000-2011)." [e6(p.1,292)] | ALS |
| Germany ALS Registry Swabia[e7,8] | The objectives of the population-based ALS registry Swabia are "to determine the incidence, prevalence and mortality of ALS in a defined geographical region in the South of Germany. […] Potential risk factors such as lifetime history of physical activity, sports, head injuries, and metabolic factors will be further investigated. […] Elucidate the underlying | ALS |

| | | |
|---|---|---|
| | pathophysiological mechanisms and to identify potential starting points for the development of new treatment approaches." [e7(p.2)] | |
| Study[e9] including 6 registries: Irish ALS Register, Scottish MND Register, Lancashire Register, the Piemonte Register, the Lombardy Register, and the Puglia Register | "This study was based on data from six prospective population-based ALS registries in three European countries (Ireland, UK and Italy) in the 2-year period 1998-1999, with a reference population of almost 24 million. These registers included the Irish ALS Register (catchment population of 3.9 million Irish residents), the Scottish MND Register (5.1 million Scottish residents), the Lancashire Register (1.6 million residents in Northwest England), the Piemonte Register (4.3 million residents in Northern Italy), the Lombardy Register (4.9 million residents in Northern Italy) and the Puglia Register (4.1 million residents in Southern Italy)." [e9(p.385-386)] | ALS |
| Italy Liguria ALS Registry (LIGALS) [e10,11] | "Assess the incidence and trends of amyotrophic lateral sclerosis (ALS) in Liguria, a north-west region of Italy, utilizing a prospective design." [e11(p.52)] | All forms of motor neuron disease. Studies cited only looked only at ALS. |
| Dutch Computer Registry of All Myopathies and Polyneuropathies (CRAMP) [e12] | "One of the incentives was to gather up-to-date nationwide epidemiological data regarding neuromuscular disorders." [e12(p.447)] | 30 neuromuscular disorders |
| Norwegian Patient Registry (NPR) [e13] | The cited study aimed to combine Norwegian Patient Registry (NPR), the Norwegian Prescription Database (NorPD), the Norwegian Cause of Death Registry (NCoDR) to determine the prevalence of ALS in Norway from 2009 to 2015. The NPR is "an administrative health register for all inpatient and outpatient admissions to Norwegian hospitals and private practice specialists with public reimbursement." [e13(p.304)] | ALS |
| Scotland Clinical Audit Research and Evaluation of Motor Neuron Disease (CARE-MND) [e14] | The aims of CARE-MND include: "1) To create a patient-centered approach to care based on recognized standards. 2) To standardize data sharing between healthcare professionals in "real-time". 3) To permit regular audit of care to drive improvements in service delivery 4) To integrate existing Scottish MND biomarker research repositories. 5) To improve patient participation in research studies including observational and clinical trials." [e14(p.243)] | ALS, or another MND subtype (PLS, PBP, PMA) |

| | | |
|---|---|---|
| Spanish National Rare Diseases Registry[e15] | "The aim of this study was to analyse the coverage of hospital discharge data and the mortality registry of La Rioja to ascertain MND cases to be included in the Spanish National Rare Diseases Registry." [e15(p.275)] | MND |
| Swedish Motor Neuron Disease Quality Registry[e16] | "Our primary aim was to construct a national quality registry for ALS patients in Sweden to improve ALS care, both including an earlier and more precise diagnosis and following care over time for patients and their families. Our secondary aim was to create a research base by prospectively following the entire ALS population in Sweden." [e16(p.528-529)] | ALS |
| **North America** | | |
| Canadian Neuromuscular Disease Registry (CNDR) [e17-19] | The CNDR aims "to improve the future for Canadians with NMD through the enablement and support of research into potential treatments. Secondary objectives include enhancing the understanding of NMD epidemiology, addressing variations in management, and facilitating clinical knowledge translation." [e18(p.699)] Over the past decade, "the registry has evolved with additional goals of improving health outcomes, pharmacoeconomic analyses, and most recently collection of post-approval real world evidence (RWE) for novel therapies." [e17(p.55)] | ALS, Duchenne Muscular Dystrophy (DMD), Myotonic Dystrophy (DM), Limb Girdle Muscular Dystrophy (LGMD), and Spinal Muscular Atrophy (SMA) |
| Neuromuscular Registry of Saguenay-Lac-Saint-Jean (SLSJ) [e20] | The Neuromuscular Registry of SLSJ, Québec, Canada was "established for epidemiological surveillance of neuromuscular disorders including amyotrophic lateral sclerosis (ALS)." The objectives of the cited study were to "analyze the ALS clinical characteristics of the SLSJ population, determine the incidence rate over time by five-year periods since 1985, and validate the Neuromuscular Registry." [e20(p.705)] | Neuromuscular disorders, including ALS. |
| Other: Integrated Neurodegenerative Disease Database (INDD) [e21] | The authors state: "It is critical that we conduct multidisciplinary patient-oriented clinical and basic science research comparatively and in a multidimensional manner to improve the understanding and treatment options for diseases such as AD, PD, FTD, ALS, and other aging-related neurodegenerative disorders. Moreover, advances in one of these disorders could accelerate the pace of advances for other such diseases. To achieve this goal, it is essential to build an overarching | ALS, Alzheimer's Disease (AD), Parkinson's Disease (PD), Frontotemporal Dementia (FTD) |

| | | |
|---|---|---|
| | integrated neurodegenerative disease (INDD) database, which includes multiple neurodegenerative disorders such as AD, PD, FTD, ALS, and other related diseases. Specifically, using this neurodegenerative disease database as a research tool, investigators would be able to obtain data across several disease groups and conduct comparative studies to elucidate distinct and common features and mechanisms of these disorders." [e21(p.e85)] | |
| Other: Massachusetts Tracking Program[e22] | "The Tracking Program was initiated in 2002 to establish a network for the ongoing collection, integration, analysis, interpretation and dissemination of data on environmental hazards, human exposure to environmental hazards, and potentially related health outcomes." [e22(p.99)] Further, the purpose of study was to explore the utility of electronic health record (EHR) data for the surveillance of ALS. | ALS |
| Other: National Administrative Healthcare Database (United States) [e23,24] | "Use several national administrative databases to estimate ALS prevalence: Medicare (parts A and B), Medicaid, Veterans Health Administration (VHA), and Veterans Benefit Administration (VBA)." [e24(p.150)] | ALS |
| Other: New Jersey (NJ) ALS Surveillance Project[e25] | The objective of the New Jersey (NJ) ALS Surveillance Project was to "evaluate the completeness of the National ALS Registry and gather reliable and timely data to better describe the incidence, prevalence and demographic characteristics of ALS among NJ residents." [e25(p.2)] | ALS |
| Other: South Carolina ALS Surveillance Pilot Project[e26,27] | "The purpose of this study was to investigate the positive predictive value and sensitivity of the ICD-10 code G12.2, which is used to identify patients who have possibly died from ALS." [e27(p.69)] | MND (unspecified), ALS, Progressive Muscular Atrophy (PMA), Progressive Bulbar Palsy (PBP), pseudobulbar palsy, Primary Lateral Sclerosis (PLS), and other motor neuron diseases. |
| Other: The State and Metropolitan Surveillance Project (United States) [e28-30] | Studies to determine the incidence and prevalence of amyotrophic lateral sclerosis (ALS). Further, "to evaluate the completeness of the National ALS Registry. Additional goals of the surveillance projects were to collect reliable and timely information regarding ALS incidence and demographic characteristics of persons with ALS in defined geographic areas." [e30(p.129)] | ALS |

| | | |
|---|---|---|
| Other: Pooled Resource Open Access ALS Clinical Trials (PRO-ACT) [e31] | "To pool data from completed amyotrophic lateral sclerosis (ALS) clinical trials and create an open-access resource to enable a greater understanding of the phenotype and biology of ALS." [e31(p.1,719)] | ALS |
| The National ALS Registry (United States) [e23,32-40] | "The main objectives of the National ALS Registry are to describe the national incidence and prevalence of ALS; describe the demographics of persons living with ALS; and examine risk factors for the disease." [e37(p.1,379)] | ALS |
| **Oceania** | | |
| Australian National Motor Neuron Disease Observational Cohort[e41] | The objectives of this cohort study were "to capture the clinical patterns, timing of key milestones and survival of patients presenting with amyotrophic lateral sclerosis/motor neuron disease (ALS/MND) within Australia" and "to determine whether ALS/MND patients could be classified into clinical phenotypes at a national level across multiple ALS/MND clinics." [e41(p.1)] Further, to help coordinate focused care and a foundation for case ascertainment for research in ALS/MND. | ALS, MND classified into clinical phenotypes (ALS, Flail Limb, PLS) |
| New Zealand Motor Neurone Disease Registry[e42] | "The NZ MND Registry was established with the aim of facilitating participation in national and international clinical trials and research, for people in New Zealand with MND. The Registry also aims to aid researchers by assisting in the planning of research. In order to do this, the Registry collects clinical and demographic data for each participant in order to be trial ready. A secondary aim is to share anonymized data in order to promote collaboration with researchers and clinicians in Australia and Asia through the Pan-Asian Consortium for Treatment and Research in ALS (PACTALS) and further afield as opportunity arises." [e42(p.8)] | Motor Neuron Diseases (MND) |

**eReferences (Table S1)**

e1. He J, Fu JY, Chen L, et al. Multicentre, prospective registry study of amyotrophic lateral sclerosis in mainland China (CHALSR): study protocol. *BMJ Open*. 2020;10(12):e042603.
e2. Shamshiri H, Fatehi F, Davoudi F, et al. Amyotrophic lateral sclerosis progression: Iran-ALS clinical registry, a multicentre study. *Amyotrophic Lateral sclerosis & Frontotemporal Degeneration*. 2015;16(7-8):506-11.
e3. Jun KY, Park J, Oh KW, et al. Epidemiology of ALS in Korea using nationwide big data. *Journal of Neurology, Neurosurgery & Psychiatry*. 2019;90(4):395-403.
e4. Kab S, Moisan F, Preux PM, Marin B, Elbaz A. Nationwide incidence of motor neuron disease using the French health insurance information system database. *Amyotrophic Lateral sclerosis & Frontotemporal Degeneration*. 2017;18(5-6):426-433.
e5. Vasta R, Boumediene F, Couratier P, et al. Validity of medico-administrative data related to amyotrophic lateral sclerosis in France: A population-based study. *Amyotrophic Lateral sclerosis & Frontotemporal Degeneration*. 2017;18(1-2):24-31.
e6. Marin B, Hamidou B, Couratier P, et al. Population-based epidemiology of amyotrophic lateral sclerosis (ALS) in an ageing Europe--the French register of ALS in Limousin (FRALim register). *European Journal of Neurology*. 2014;21(10):1292-300, e78.
e7. Nagel G, Unal H, Rosenbohm A, et al. Implementation of a population-based epidemiological rare disease registry: Study protocol of the amyotrophic lateral sclerosis (ALS) - registry Swabia. *BMC Neurology*. 2013;13 (no pagination)
e8. Uenal H, Rosenbohm A, Kufeldt J, et al. Incidence and geographical variation of amyotrophic lateral sclerosis (ALS) in Southern Germany--completeness of the ALS registry Swabia. *PLoS ONE [Electronic Resource]*. 2014;9(4):e93932.
e9. Logroscino G, Traynor BJ, Hardiman O, et al. Incidence of amyotrophic lateral sclerosis in Europe. *Journal of Neurology, Neurosurgery & Psychiatry*. 2010;81(4):385-90.
e10. Scialo C, Novi G, Bandettini di Poggio M, et al. Clinical epidemiology of amyotrophic lateral sclerosis in Liguria, Italy: An update of LIGALS register. *Amyotrophic Lateral sclerosis & Frontotemporal Degeneration*. 2016;17(7-8):535-542.
e11. Bandettini di Poggio M, Sormani MP, Truffelli R, et al. Clinical epidemiology of ALS in Liguria, Italy. *Amyotrophic Lateral sclerosis & Frontotemporal Degeneration*. 2013;14(1):52-7.
e12. Deenen JC, van Doorn PA, Faber CG, et al. The epidemiology of neuromuscular disorders: Age at onset and gender in the Netherlands. *Neuromuscular Disorders*. 2016;26(7):447-52.
e13. Nakken O, Lindstrom JC, Tysnes OB, Holmoy T. Assessing amyotrophic lateral sclerosis prevalence in Norway from 2009 to 2015 from compulsory nationwide health registers. *Amyotrophic Lateral sclerosis & Frontotemporal Degeneration*. 2018;19(3-4):303-310.
e14. Leighton D, Newton J, Colville S, et al. Clinical audit research and evaluation of motor neuron disease (CARE-MND): a national electronic platform for prospective, longitudinal monitoring of MND in Scotland. *Amyotrophic Lateral sclerosis & Frontotemporal Degeneration*. 2019;20(3-4):242-250.
e15. Ruiz E, Ramalle-Gomara E, Quinones C, Spain RDR Working Group. Record linkage between hospital discharges and mortality registries for motor neuron disease case ascertainment for the Spanish National Rare Diseases Registry. *Amyotrophic Lateral sclerosis & Frontotemporal Degeneration*. 2014;15(3-4):275-8.

e16. Longinetti E, Regodon Wallin A, Samuelsson K, et al. The Swedish motor neuron disease quality registry. *Amyotrophic Lateral sclerosis & Frontotemporal Degeneration*. 2018;19(7-8):528-537.
e17. Hodgkinson V, Lounsberry J, M'Dahoma S, et al. The Canadian Neuromuscular Disease Registry 2010-2019: A Decade of Facilitating Clinical Research Througha Nationwide, Pan-NeuromuscularDisease Registry. *Journal of neuromuscular diseases*. 2021;8(1):53-61.
e18. Korngut L, Campbell C, Johnston M, et al. The CNDR: Collaborating to translate new therapies for Canadians. *Canadian Journal of Neurological Sciences*. 2013;40(5):698-704.
e19. Korngut L, Genge A, Johnston M, et al. Establishing a Canadian registry of patients with amyotrophic lateral sclerosis. *Canadian Journal of Neurological Sciences*. 2013;40(1):29-35.
e20. Lareau-Trudel E, Fortin E, Gauthier M, Lavoie S, Morissette E, Mathieu J. Epidemiological surveillance of amyotrophic lateral sclerosis in Saguenay region. *Canadian Journal of Neurological Sciences*. 2013;40(5):705-9.
e21. Xie SX, Baek Y, Grossman M, et al. Building an integrated neurodegenerative disease database at an academic health center. *Alzheimer's & Dementia*. 2011;7(4):e84-93.
e22. Namulanda G, Qualters J, Vaidyanathan A, et al. Electronic health record case studies to advance environmental public health tracking. *Journal of Biomedical Informatics*. 2018;79:98-104.
e23. Kaye WE, Sanchez M, Wu J. Feasibility of creating a National ALS Registry using administrative data in the United States. *Amyotrophic Lateral sclerosis & Frontotemporal Degeneration*. 2014;15(5-6):433-9.
e24. Nelson LM, Topol B, Kaye W, et al. Estimation of the Prevalence of Amyotrophic Lateral Sclerosis in the United States Using National Administrative Healthcare Data from 2002 to 2004 and Capture-Recapture Methodology. *Neuroepidemiology*. 2018;51(3-4):149-157.
e25. Jordan H, Fagliano J, Rechtman L, Lefkowitz D, Kaye W. Population-based surveillance of amyotrophic lateral sclerosis in New Jersey, 2009-2011. *Neuroepidemiology*. 2014;43(1):49-56.
e26. Stickler DE, Royer JA, Hardin JW. Validity of hospital discharge data for identifying cases of amyotrophic lateral sclerosis. *Muscle and Nerve*. 2011;44(5):814-815.
e27. Stickler DE, Royer JA, Hardin JW. Accuracy and usefulness of ICD-10 death certificate coding for the identification of patients with ALS: results from the South Carolina ALS Surveillance Pilot Project. *Amyotrophic Lateral Sclerosis*. 2012;13(1):69-73.
e28. Freer C, Hylton T, Jordan HM, Kaye WE, Singh S, Huang Y. Results of Florida's Amyotrophic Lateral Sclerosis Surveillance Project, 2009-2011. *BMJ Open*. 2015;5(4):e007359.
e29. Jordan H, Rechtman L, Wagner L, Kaye WE. Amyotrophic lateral sclerosis surveillance in Baltimore and Philadelphia. *Muscle & Nerve*. 2015;51(6):815-21.
e30. Wagner L, Rechtman L, Jordan H, et al. State and metropolitan area-based amyotrophic lateral sclerosis (ALS) surveillance. *Amyotrophic Lateral Sclerosis and Frontotemporal Degeneration*. 2016;17(1-2):128-134.
e31. Atassi N, Berry J, Shui A, et al. The PRO-ACT database Design, initial analyses, and predictive features. *Neurology*. 2014;83(19):1719-1725.
e32. Antao VC, Horton DK. The National Amyotrophic Lateral Sclerosis (ALS) Registry. *Journal of Environmental Health*. 2012;75(1):28-30.
e33. Benatar M, Wuu J, Usher S, Ward K. Preparing for a U.S. National ALS Registry: Lessons from a pilot project in the State of Georgia. *Amyotrophic Lateral Sclerosis*. 2011;12(2):130-5.

e34. Bryan L, Kaye W, Antao V, Mehta P, Muravov O, Horton DK. Preliminary Results of National Amyotrophic Lateral Sclerosis (ALS) Registry Risk Factor Survey Data. *PLoS ONE [Electronic Resource]*. 2016;11(4):e0153683.
e35. Kaye WE, Wagner L, Wu R, Mehta P. Evaluating the completeness of the national ALS registry, United States. *Amyotrophic Lateral sclerosis & Frontotemporal Degeneration*. 2018;19(1-2):112-117.
e36. Malek AM, Stickler DE, Antao VC, Horton DK. The National ALS Registry: a recruitment tool for research. *Muscle & Nerve*. 2014;50(5):830-4.
e37. Mehta P, Horton DK, Kasarskis EJ, et al. CDC Grand Rounds: National Amyotrophic Lateral Sclerosis (ALS) Registry Impact, Challenges, and Future Directions. *Mmwr*. 2017;Morbidity and mortality weekly report. 66(50):1379-1382.
e38. Raymond J, Oskarsson B, Mehta P, Horton K. Clinical characteristics of a large cohort of US participants enrolled in the National Amyotrophic Lateral Sclerosis (ALS) Registry, 2010-2015. *Amyotrophic Lateral sclerosis & Frontotemporal Degeneration*. 2019;20(5-6):413-420.
e39. Valle J, Roberts E, Paulukonis S, Collins N, English P, Kaye W. Epidemiology and surveillance of amyotrophic lateral sclerosis in two large metropolitan areas in California. *Amyotrophic Lateral sclerosis & Frontotemporal Degeneration*. 2015;16(3-4):209-15.
e40. Wittie M, Nelson LM, Usher S, Ward K, Benatar M. Utility of capture-recapture methodology to assess completeness of amyotrophic lateral sclerosis case ascertainment. *Neuroepidemiology*. 2014;40(2):133-141.
e41. Talman P, Duong T, Vucic S, et al. Identification and outcomes of clinical phenotypes in amyotrophic lateral sclerosis/motor neuron disease: Australian National Motor Neuron Disease observational cohort. *BMJ Open*. 2016;6(9):e012054.
e42. Walker KL, Rodrigues MJ, Watson B, et al. Establishment and 12-month progress of the New Zealand Motor Neurone Disease Registry. *Journal of Clinical Neuroscience*. 2019;60:7-11.
